# The U.S. COVID-19 Trends and Impact Survey, 2020-2021: Continuous real-time measurement of COVID-19 symptoms, risks, protective behaviors, testing and vaccination

**DOI:** 10.1101/2021.07.24.21261076

**Authors:** Joshua A. Salomon, Alex Reinhart, Alyssa Bilinski, Eu Jing Chua, Wichada La Motte-Kerr, Minttu M. Rönn, Marissa Reitsma, Katherine Ann Morris, Sarah LaRocca, Tamar Farag, Frauke Kreuter, Roni Rosenfeld, Ryan J. Tibshirani

## Abstract

The U.S. COVID-19 Trends and Impact Survey (CTIS) is a large, cross-sectional, Internet-based survey that has operated continuously since April 6, 2020. By inviting a random sample of Facebook active users each day, CTIS collects information about COVID-19 symptoms, risks, mitigating behaviors, mental health, testing, vaccination, and other key priorities. The large scale of the survey – over 20 million responses in its first year of operation – allows tracking of trends over short timescales and allows comparisons at fine demographic and geographic detail. The survey has been repeatedly revised to respond to emerging public health priorities. In this paper, we describe the survey methods and content and give examples of CTIS results that illuminate key patterns and trends and help answer high-priority policy questions relevant to the COVID-19 epidemic and response. These results demonstrate how large online surveys can provide continuous, real-time indicators of important outcomes that are not subject to public health reporting delays and backlogs. The CTIS offers high value as a supplement to official reporting data by supplying essential information about behaviors, attitudes toward policy and preventive measures, economic impacts, and other topics not reported in public health surveillance systems.

**Significance statement:** The U.S. COVID-19 Trends and Impact Survey (CTIS) has operated continuously since April 6, 2020, collecting over 20 million responses. The largest public health survey ever conducted in the United States, CTIS was designed to facilitate detailed demographic and geographic analyses, track trends over time, and accommodate rapid response to emerging priorities. Using examples of CTIS results illuminating trends in symptoms, risks, mitigating behaviors, testing and vaccination in relation to evolving high-priority policy questions over 12 months of the pandemic, we illustrate the value of online surveys for tracking patterns and trends in COVID outcomes as an adjunct to official reporting, and showcase unique insights that would not be visible through traditional public health reporting.

## Introduction

During 2020, the coronavirus disease 2019 (COVID-19) pandemic precipitated the need for new public health surveillance to inform urgent policy decisions. Effective pandemic policy making requires information on a broad array of indicators including local morbidity and mortality, preventive behaviors, healthcare capacity, and economic impacts. Given the critical importance of COVID-19 trends for policy, health departments set up routine public reporting systems for tracking cases, deaths, testing, and hospitalizations (1). However, supplemental data can both augment official reporting, for example by providing additional indicators of COVID-19 prevalence not subject to reporting delays and backlogs, and supply complementary information about public behavior, attitudes toward policy and preventive measures, mental health, economic impacts, and other items not observed in public health surveillance systems.

A number of efforts have used surveys to provide supplemental surveillance data. For example, symptom-tracking smartphone apps invite users to self-report symptoms, in some cases encouraging repeated participation to enable longitudinal tracking (2-4). Other surveys have addressed broader impacts of the pandemic, such as economic consequences (5). In this paper, we present findings from the Delphi Group at Carnegie Mellon University U.S. COVID-19 Trends and Impact Survey (US CTIS), in partnership with Facebook, which has operated continuously since April 6, 2020 and collected over 20 million responses. (An international version of the survey is described in a companion paper in this theme issue.) A random sample of Facebook active users are invited each day to complete a questionnaire comprising survey items on symptoms, COVID testing, social distancing, vaccination, schooling, mental health, and economic security. Results are aggregated and made publicly available, and microdata are available under institutional data use agreement, in both cases with less than three days of lag. These data provide information at a level of geographic and temporal detail that can supply essential inputs into short-term decision-making and longer-term strategic planning. The survey instrument has been updated frequently to incorporate new policy-relevant topics. The largest public health survey ever conducted in the United States, CTIS is designed to facilitate detailed demographic and geographic analyses, to track trends over time, and to accommodate rapid response to emerging priorities (6).

In this study, we first compare COVID-19 indicators from CTIS with publicly-reported case, hospitalization and mortality data between April 2020 and April 2021. Despite potential limitations of our Internet-based sample and the voluntary nature of the survey, we demonstrate high correspondence between the two, with CTIS less affected by holiday-related reporting anomalies. Second, we examine patterns and trends in symptoms, risks, mitigating behaviors, testing and vaccination in US states and localities in relation to evolving high-priority policy questions over 12 months of the pandemic. The findings illustrate the value of online surveys for tracking patterns and trends in COVID-related outcomes as an adjunct to official reporting, while also showcasing insights that are only possible through a large-scale survey effort.

## Methods

### Sampling and recruitment

The U.S. COVID-19 Trends and Impact Survey launched on April 6, 2020 and has run continuously since that time, with an average of more than 350,000 people participating each week over the first year of operation. The survey is implemented by the Delphi Group at Carnegie Mellon University (CMU), with participants recruited via the Facebook platform. Every day, Facebook invites a new sample of active users ages 18 years or older to participate in the survey. Facebook uses stratified random sampling within US states to randomly select a sample of its users to see the survey invitation at the top of their News Feed. Users who click on the invitation are taken to the CMU-administered survey hosted on Qualtrics. To ensure privacy, Facebook does not see any individual survey response during or after the data collection. The survey is available in English, Spanish, Brazilian Portuguese, Vietnamese, French, and simplified Chinese.

### Survey design

The survey instrument was deployed in 10 waves from launch through April 5, 2021, with contents of each survey version summarized in Table 1. Revisions are ongoing as new public health needs arise. A number of core items have been included consistently across all survey versions, including questions about symptoms, contacts and demographics. Key additions include items on mask wearing and occupation, added in September 2020, seasonal flu vaccination and schooling, added in November 2020, and COVID-19 vaccination, added in December 2020. As of April 5, 2021, the range of survey items spanned the following broad categories: household and individual symptoms, common comorbidities, contact patterns and mitigating behaviors, testing and diagnosis, worry and financial impact, schooling, vaccination, and demographics. Full versions of all survey instruments can be found at https://cmu-delphi.github.io/delphi-epidata/symptom-survey/coding.html.

**Table 1:**
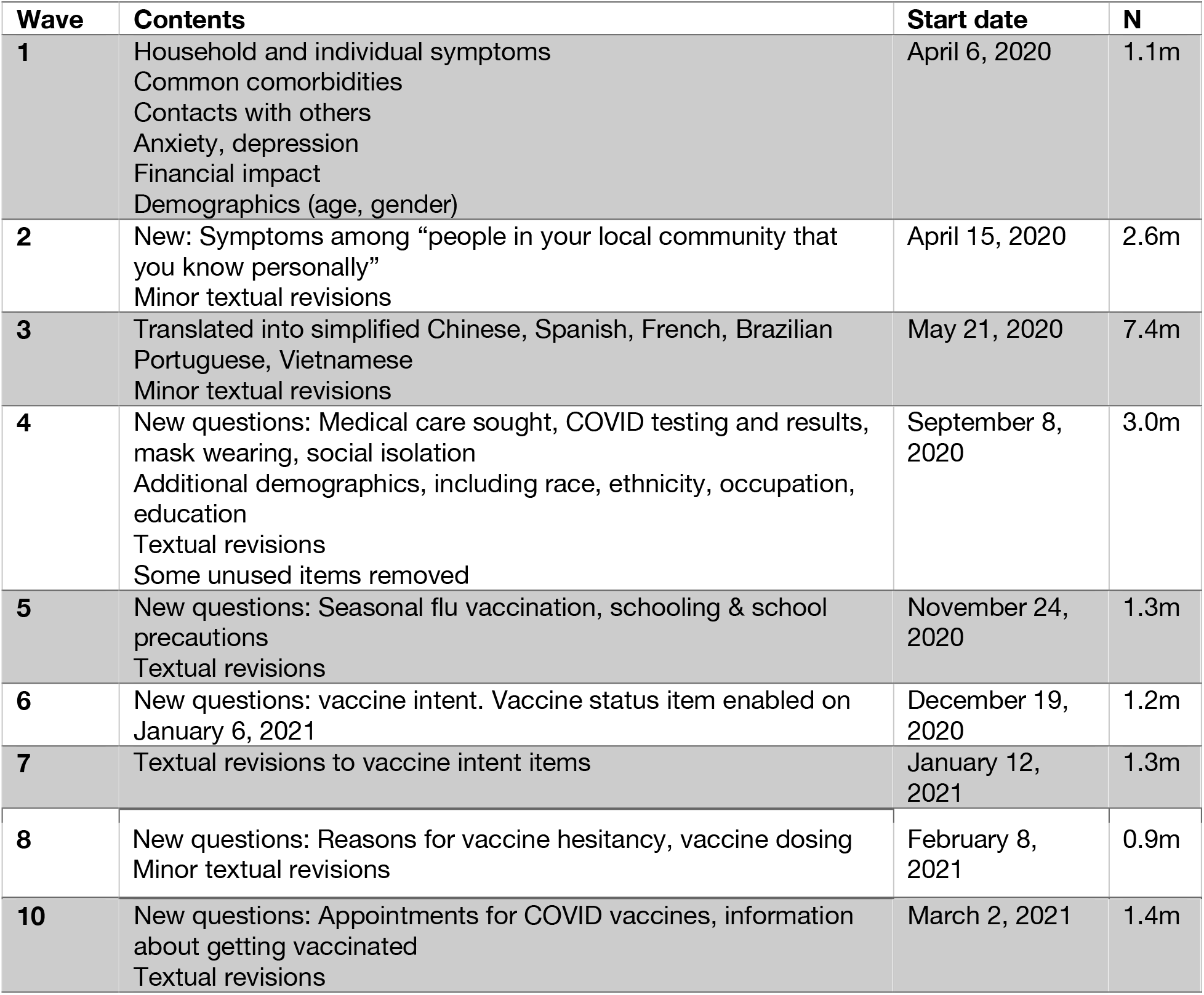
Summary of survey waves deployed between April 6, 2020 and April 5, 2021.

### Weighting

Analytic weights have been developed to adjust for differences between Facebook users and the United States population, and to adjust for biases related to coverage and non-response (7). When Facebook links users to the survey, it generates a random unique identifier that is passed to CMU. For users who complete the survey, CMU returns the corresponding identifiers to Facebook, which then calculates analytic weights in two steps:

1. To adjust for non-response bias, Facebook calculates the inverse probability that sampled users complete the survey using their age, gender, and geographical variables, as reported on their Facebook profiles, as well as other characteristics known to correlate with non-response. The inverse probabilities are then used to create weights for responses, after which the survey sample reflects the active adult user population on Facebook.
2. To adjust for coverage bias, Facebook post-stratifies the weights created in the first step so that the distribution of age, gender and state or territory of residence in the survey sample reflects that of the general population.

The weight value does not identify the survey respondent. The weight value for an individual is scaled to approximate the number of people in the adult population represented by that individual based on age, gender, location, and date. Facebook passes these weights to CMU. CMU cannot use these weights to identify specific Facebook users, and Facebook never receives individual survey responses and cannot link them to specific users.

### Analysis

In this study we examined a range of different outcomes measured in the CTIS over the period April 6, 2020 to April 5, 2021. Characteristics of the study sample were compared to data from the American Community Survey 2019 supplemental estimates. We evaluated reported symptoms and symptom patterns in comparison to surveillance data on confirmed COVID-19 hospitalizations from the Department of Health and Human Services (8) and reported COVID-19 cases and mortality aggregated by the Johns Hopkins University Center for Systems Science and Engineering (9).

We defined a condition called *COVID-like illness* or *CLI*, which comprised reporting a fever of at least 100° F, along with cough, shortness of breath, or difficulty breathing, in line with a working definition of CLI used for syndromic surveillance purposes beginning in early 2020. A second indicator was constructed based on responses to an item on the survey that asks whether respondents know someone personally in their community who is ill with COVID-like symptoms, which we call *CLI-in-Community*.

Some results were stratified by county groupings defined in references to the CDC Social Vulnerability Index (10), which is a composite measure constructed based on 15 social variables measured at the census tract level. We have analyzed CTIS data using demographic groupings that include age, race/ethnicity using categories consistent with National Center for Health Statistics, geographic divisions including Census region, Census division, state and county. For measures of vaccination acceptance, we have pooled results over the period March 1, 2021 to April 5, 2021 and displayed results only for those counties with at least 50 responses recorded over that period.

### Data Availability

De-identified individual participant data are available to academic and nonprofit researchers under a Data Use Agreement that protects the confidentiality of respondents. County- and state-level aggregates of key variables are publicly available in the COVIDcast API, described in detail in a companion paper, and are presented in an interactive online dashboard (https://delphi.cmu.edu/covidcast/survey-results). Demographic breakdowns of key variables over time are available for public download at https://cmu-delphi.github.io/delphi-epidata/symptom-survey/contingency-tables.html.

The study was approved by the Carnegie Mellon University Institutional Review Board, under protocol STUDY2020_00000162.

## Results

### Characteristics of the study sample

As of April 5, 2021, a total of 20.2 million responses had been collected in the United States COVID Trends and Impact Survey. Table 2 summarizes characteristics of the survey respondents. Compared to the weighted sample, the unweighted sample had a higher proportion of women (66% vs. 52%) and a slightly higher proportion of respondents between ages 25 and 64 years (72% vs 68%). Household size and prevalence of at least one comorbidity were similar in the unweighted and weighted samples.

**Table 2:**
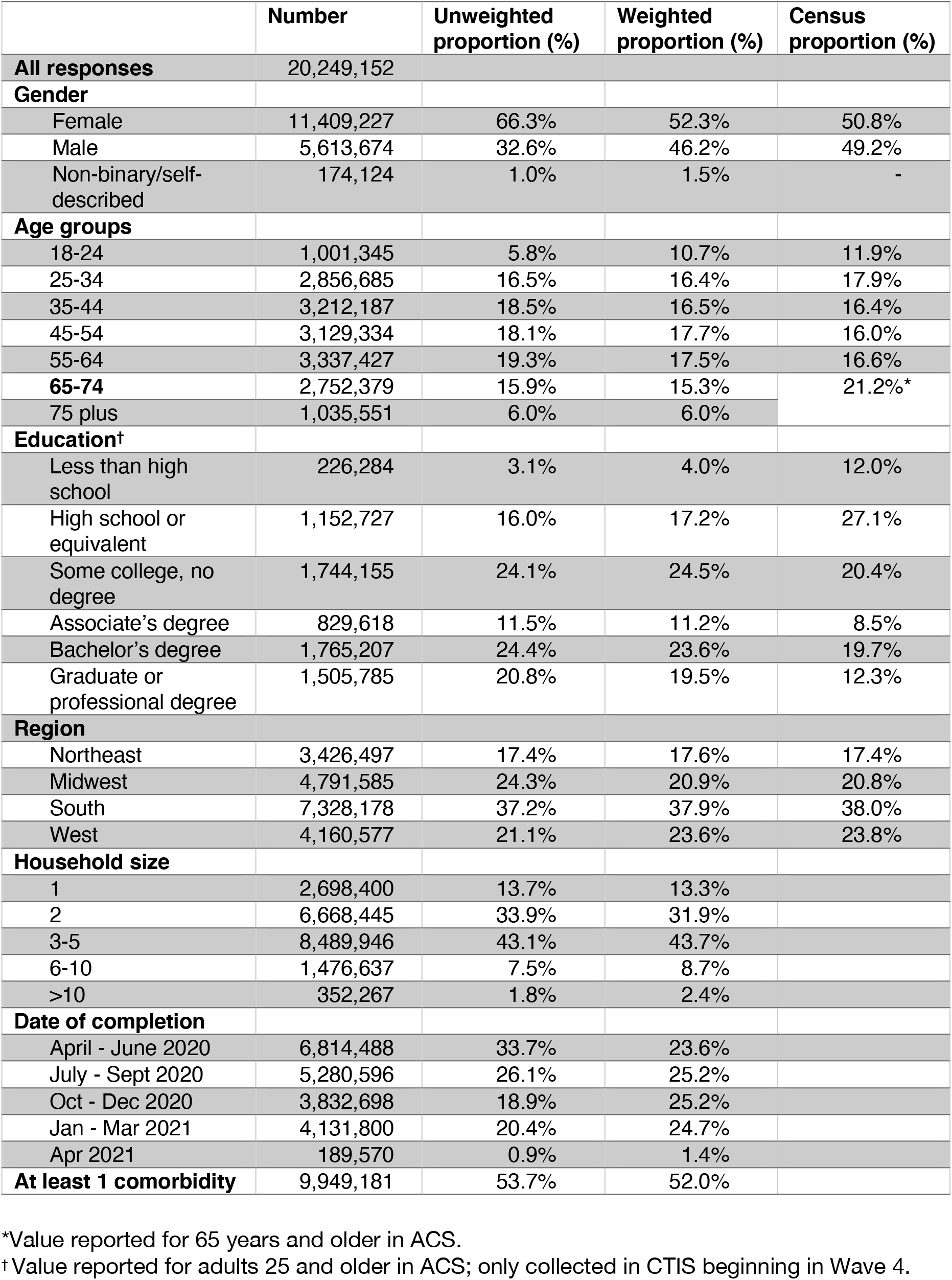
Characteristics of the study sample, compared to 2019 American Community Survey supplemental estimates.

Compared to 2019 American Community Survey supplemental estimates, the weighted survey sample slightly overrepresented women, but had a broadly comparable age and geographic distribution. The weighted sample included a larger proportion of respondents with greater than a high school education, and a much smaller proportion with less than a high school education, suggesting the presence of a sampling or response bias correlated with education. This bias was consistent since education was added to the survey instrument. As the weights provided by Facebook do not account for education, the weighting did not correct this bias.

### COVID-19 symptoms and diagnoses

A large fraction of daily respondents reported new or unusual symptoms consistent with COVID-19 (Figure 1). The most common single new or unusual symptom reported among all respondents was “tiredness or exhaustion,” with a prevalence of 3.9% among all reports in Waves 4-10. Patterns of symptom reports were notably different among those who reported testing positive for COVID-19 compared to all other respondents, including a significantly higher probability of reporting loss of smell or taste (34% compared to 1.2%).

**Figure 1.**
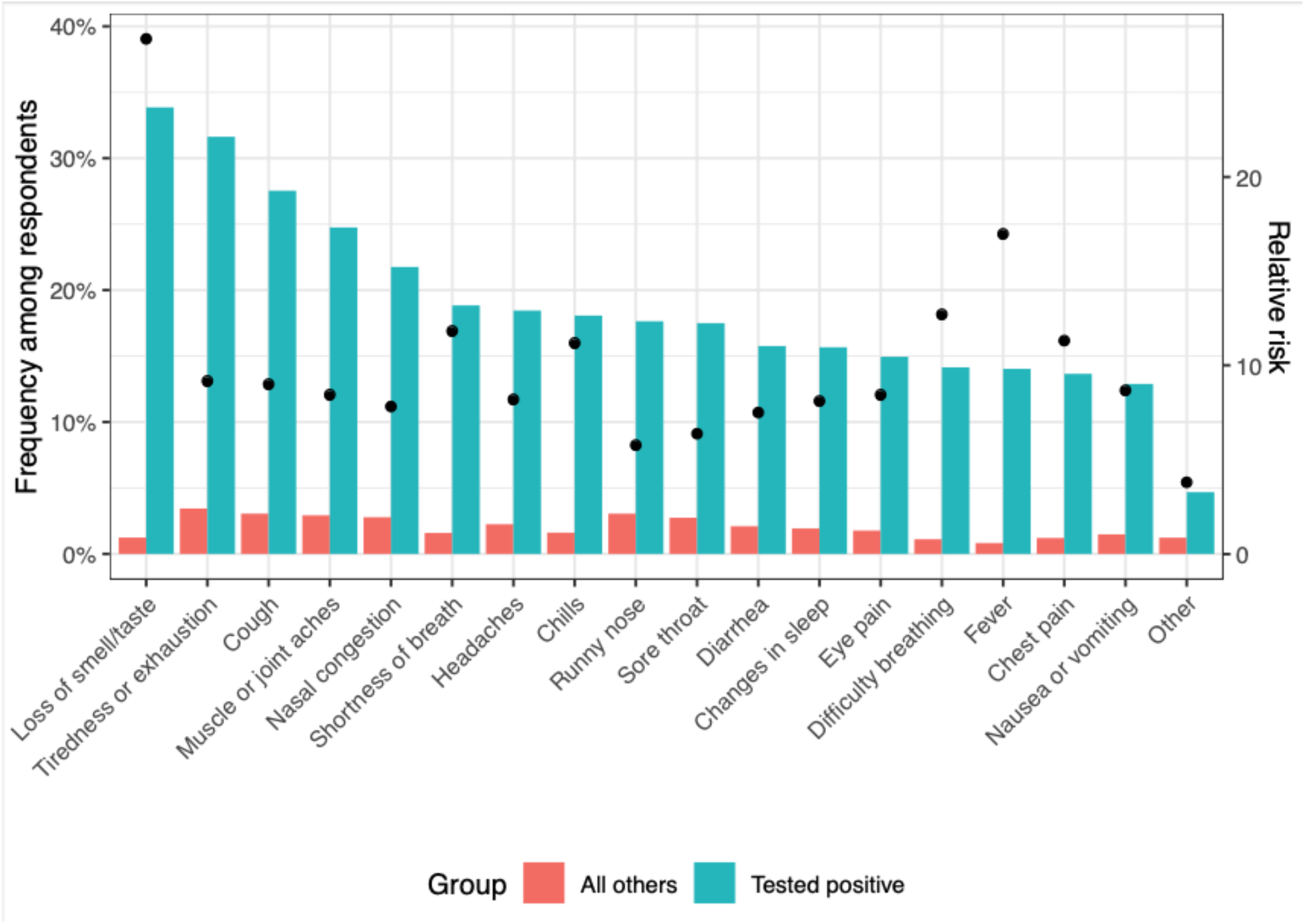
Frequency of reported new or unusual symptoms, pooled over respondents to the COVID Trends and Impact Survey, September 8, 2020 to April 5, 2021. Respondents are grouped by whether they indicated they tested positive in the past 14 days. Dots indicate the ratio of frequency among those who tested positive compared to all others.

Figure 2 compares time series for three indicators from the CTIS: reported anosmia, CLI and CLI-in-community, against the three main surveillance indicators that have been used to monitor trends in the epidemic: confirmed cases, hospitalizations and deaths, stratified by Census region. Over the period April 6, 2020 to April 5, 2021, the three survey indicators tracked both broad temporal trends and regional patterns in the surveillance indicators, and several notable features are evident in the comparison. First, the survey-based indicators were less susceptible to daily fluctuations and reporting anomalies that appeared in cases and deaths, including abrupt discontinuities around certain holiday periods. Second, trends and patterns in anosmia were similar to patterns in CLI, and the anosmia series provided a closer match than the other two survey indicators to the trends and patterns observed in COVID-19 hospitalizations, mirroring temporal peaks and ordering of levels across regions over different waves of the epidemic. Third, CLI-in-community, with the highest levels of prevalence, provided the most temporally stable signals while also expressing broad differences over time and space that were generally similar to those in other indicators. In a companion paper, we performed extensive correlation analyses between reported cases and various auxiliary indicators including the survey-based CLI and CLI-in-community signals, showing strong correlations between cases and these two survey signals during much of the pandemic.

**Figure 2.**
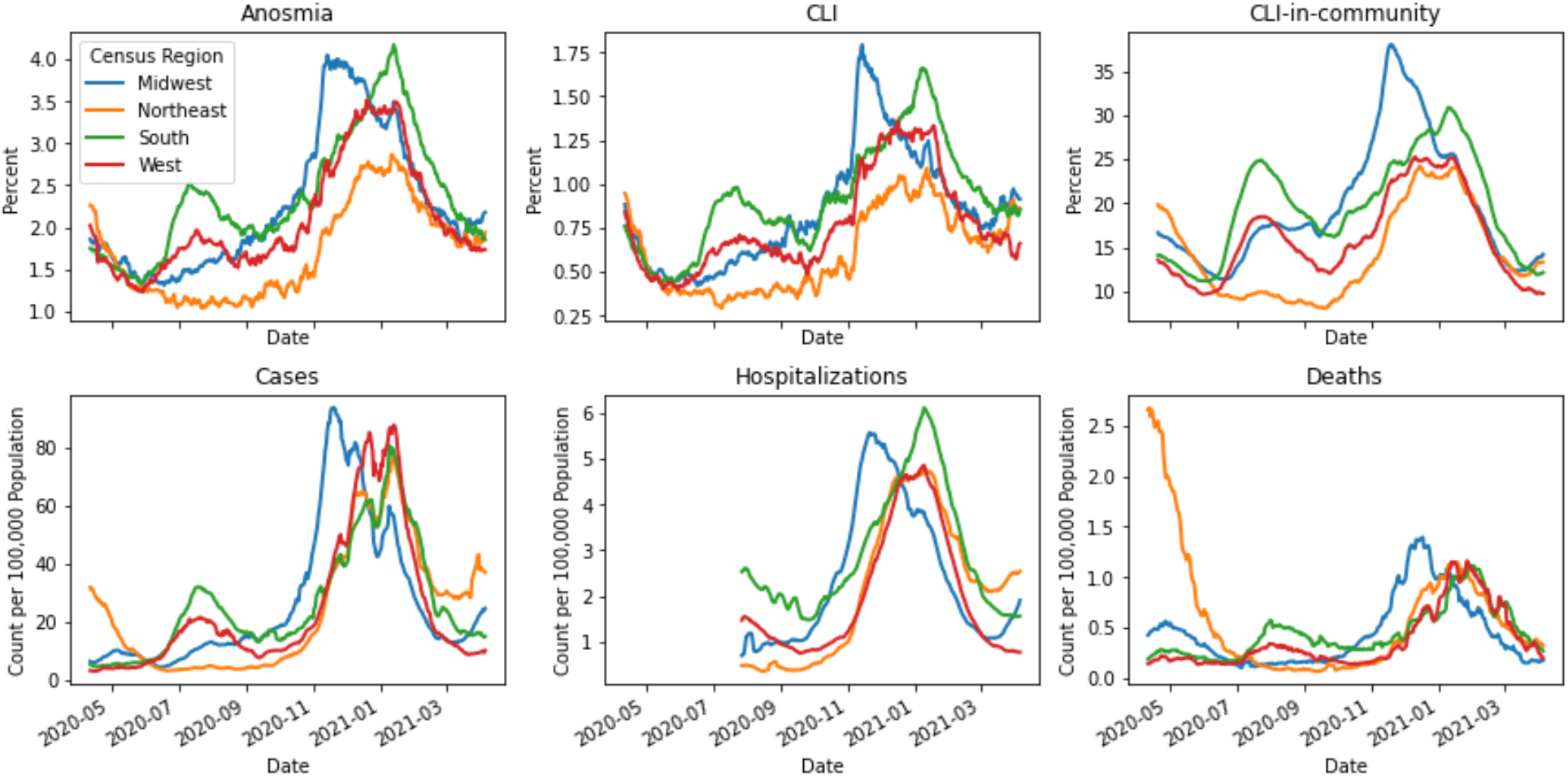
Trends in anosmia, COVID-like illness (CLI), CLI-in-community, confirmed cases, hospitalizations and deaths, by Census region, April 6, 2020 to April 5, 2021.

The CTIS includes questions about testing and diagnosis, which since September 8, 2020 have been asked of all respondents. Figure 3 compares weekly CTIS estimates of the proportion of adults reporting that they have ever had a positive test for COVID-19 against cumulative diagnoses from surveillance reports by state in the same week. State surveillance reports were adjusted, using American Community Survey 5-year population estimates and CDC line-level demographic data on confirmed COVID-19 cases, to produce estimated diagnosis rates among the state’s population over age 18. As of April 5, 2021, reported diagnoses in the survey ranged from 3.1% in Hawaii to 19% in Idaho, and the correlation between survey reported diagnoses and surveillance reports at state level was 0.83, indicating strong convergent validity.

**Figure 3.**
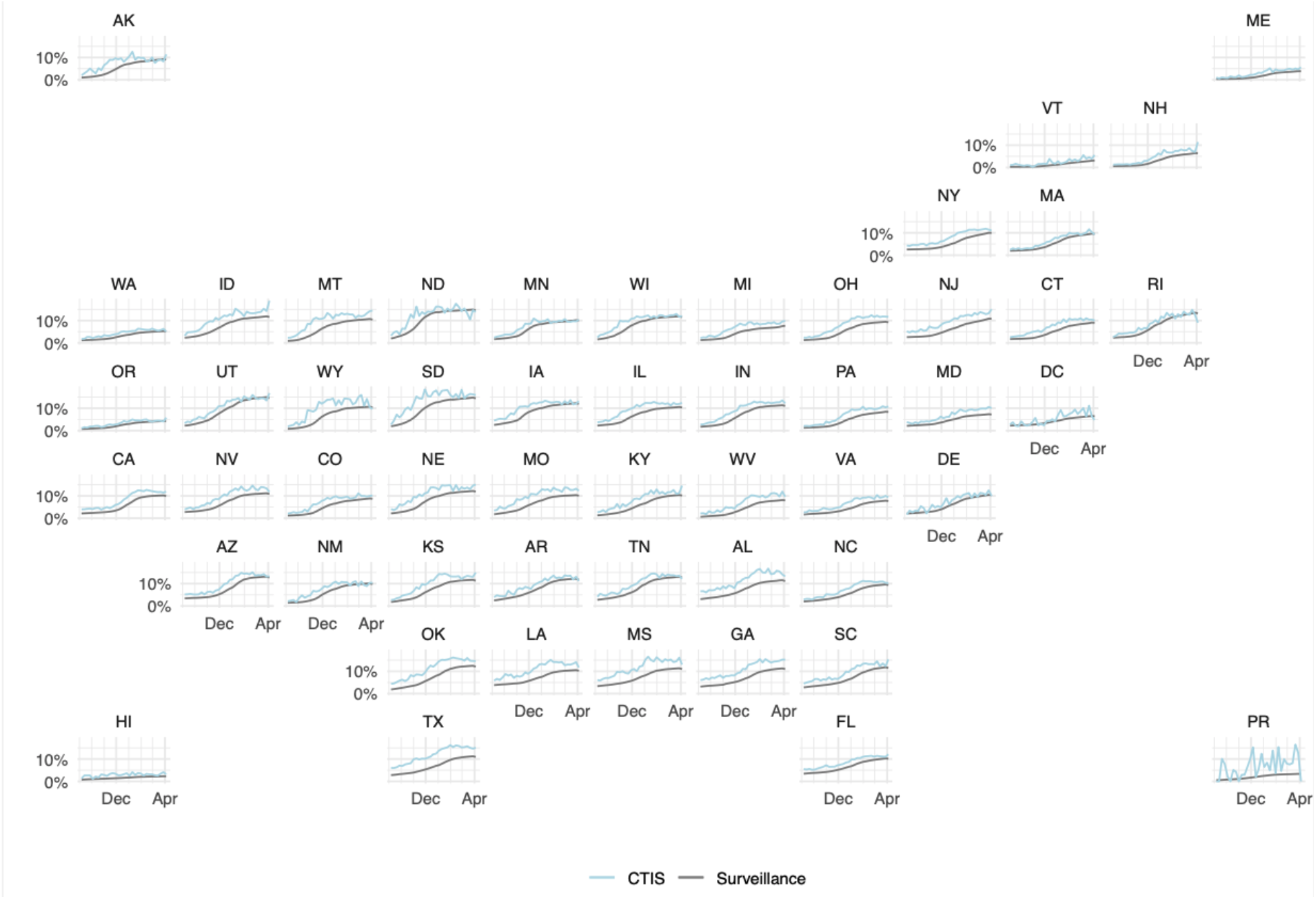
Comparison of proportion of respondents in COVID Trends and Impact Survey reporting ever having tested positive for COVID-19 and cumulative proportion of adult population with confirmed COVID diagnosis, by state, September 8, 2020 through April 5, 2021

### Transmission risk by individual characteristics

Since Wave 4, the CTIS has included questions about occupation, which can offer valuable insights into exposures among essential workers and also supply signals of where transmission may be concentrated. Using responses from January 2021, we examined the probability of reporting a positive COVID-19 test across different occupation categories, as well as the probability of reporting working outside the home while having symptoms consistent with COVID-19 (Table 3). Substantial heterogeneity appeared across broad groups of occupation. The large proportion of people reporting never having been tested indicates the limitations of passive surveillance. Combining questions on symptoms, testing and working into a single indicator, we examined the fraction of people who reported both working outside the home and currently having atypical symptoms; results ranged from more than 15% for respondents in food preparation and serving related occupations, to 4% of those in arts, design, entertainment, sports, and media.

**Table 3:** Reporting testing, symptoms, and working outside the home, by reported occupation category, in January 2021.

| **Occupation group** | **% tested positive** | **% working with symptoms** | **% working outside and never tested** | **N** |
| --- | --- | --- | --- | --- |
| Arts, design, entertainment, sports, and media | 7.2 | 4.0 | 17.1 | 20,585 |
| Building and grounds cleaning and maintenance | 11.9 | 10.8 | 45.8 | 12,528 |
| Community and social service | 13.1 | 9.0 | 23.3 | 28,223 |
| Construction and extraction | 11.8 | 11.3 | 47.7 | 10,528 |
| Education, training, and library | 10.5 | 6.8 | 24.0 | 72,098 |
| Food preparation and serving related | 13.6 | 15.5 | 39.4 | 30,817 |
| Healthcare practitioners and technicians | 15.1 | 9.6 | 25.4 | 70,793 |
| Healthcare support | 15.2 | 9.2 | 22.8 | 45,084 |
| Installation, maintenance, and repair | 10.5 | 10.9 | 49.2 | 15,511 |
| Office and administrative support | 11.0 | 6.2 | 24.4 | 84,285 |
| Other | 9.9 | 6.5 | 25.9 | 165,719 |
| Personal care and service | 12.1 | 9.2 | 32.6 | 15,115 |
| Production | 14.1 | 12.6 | 42.1 | 23,149 |
| Protective service | 14.6 | 11.8 | 33.5 | 8,314 |
| Sales and related | 11.7 | 10.8 | 36.1 | 63,066 |
| Transportation and material moving | 11.2 | 10.5 | 48.3 | 23,013 |

### Mitigating behaviors and policy analysis

A core set of questions included since the launch of the survey has addressed contacts and preventive behaviors. The survey has been amended over time to augment these, with addition of questions on mask use and specific high-risk behaviors in September 2020. In the context of recurrent surges in COVID-19 around the country over the course of 2020 and 2021, these items have illuminated how contacts and mitigating behaviors can shift in response to changes in local COVID-19 risk, sometimes preceding policy changes. For example, Figure 4 shows selected variables relating to contacts and preventive behaviors over the period September 2020 to April 2021. Responses indicate sharp increases in risk-reducing behaviors during November and December as cases surged – including reduced contacts, increased use of masks, and reduced use of public transit – followed by relaxation of mitigating behaviors over the period January to April 2021 as cases fell.

**Figure 4.**
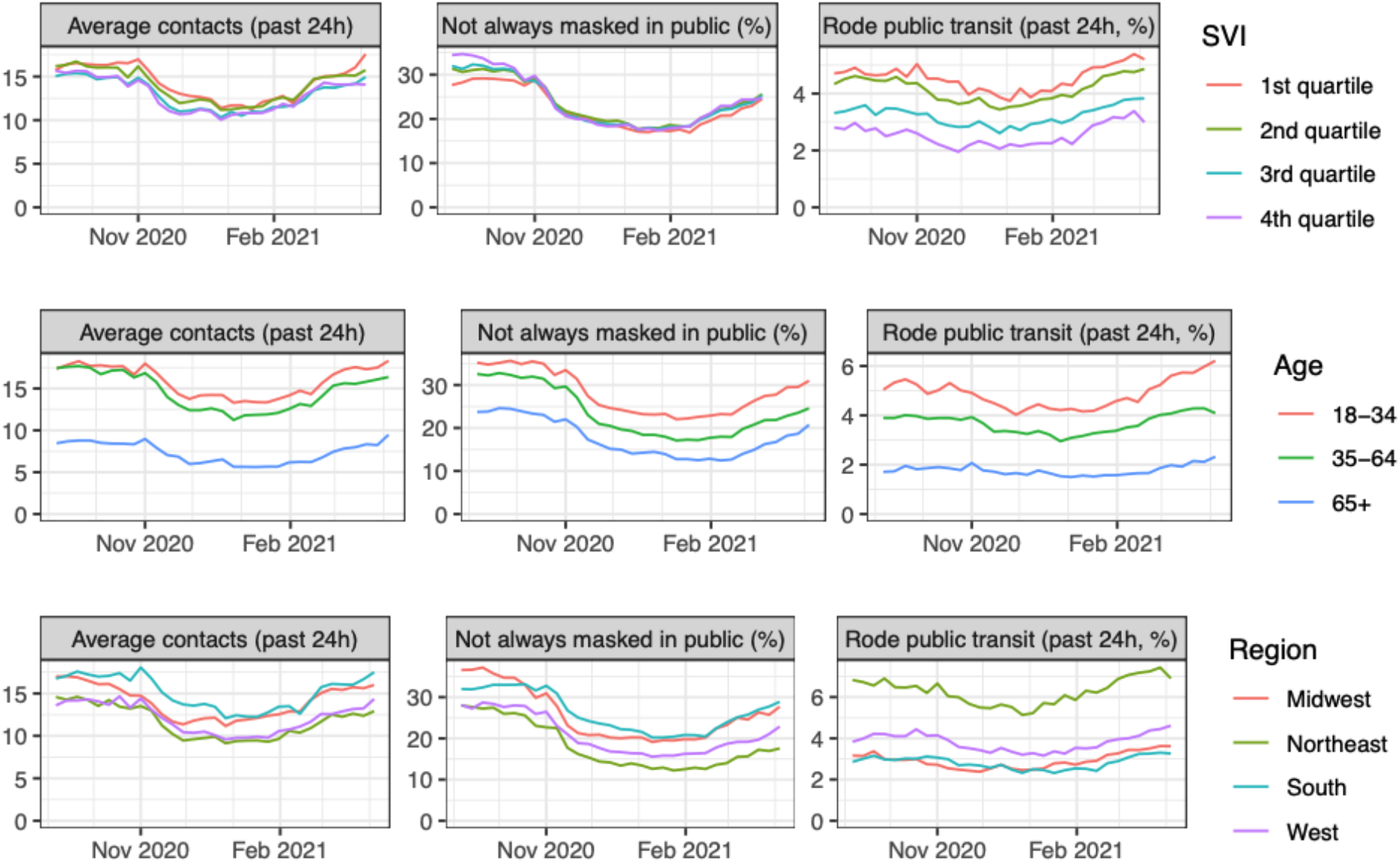
Contacts, mask use and use of public transport, by (a) quartile of counties grouped by the CDC Social Vulnerability Index, (b) age group and (c) Census region, September 8, 2020 to April 5, 2021

Individual-level data allow for geographically-detailed analysis that can also be disaggregated by demographic features. In Figure 4, results are stratified in three different ways to illustrate this: by quartiles of the CDC Social Vulnerability Index (SVI), by age and by US Census region. There were minimal differences across counties grouped by SVI in reported mask use, moderately higher contacts among those living in more vulnerable communities, and substantially higher use of public transit in more vulnerable counties. The second row shows age differences, which indicate a pronounced gradient of higher risk mitigation among older respondents, especially with respect to reduced contacts. The third row describes regional patterns that vary across indicators, with higher contacts and lower mask use in the South and Midwest regions compared to the Northeast and West, but greater use of public transit in the Northeast and West compared to South and Midwest.

### Vaccination and vaccine acceptance

Since December 19, 2020, the CTIS has included questions on vaccination intent, and since January 6, 2021, the survey has asked about vaccination status. The combination of geographic and demographic resolution in the survey allows a uniquely detailed view on vaccination acceptance and hesitancy across different US population groups. Figure 5A displays results by age group, race/ethnicity, gender, and Census region, pointing to high levels of acceptance among older respondents in all categories, but lower and more variable results at younger ages. (Respondents may identify as non-binary or self-describe their gender, but this group was typically too small to break out and report reliable hesitancy estimates by region.) Figure 5B shows the percentage of respondents indicating that they would probably not or definitely not get vaccinated across US counties, indicating regional patterns but also high variability across counties within a given state. As the vaccination campaign slows across the country, high resolution information on vaccine acceptance can inform policies that aim to increase uptake toward the goal of high levels of population immunity against COVID-19.

**Figure 5.**
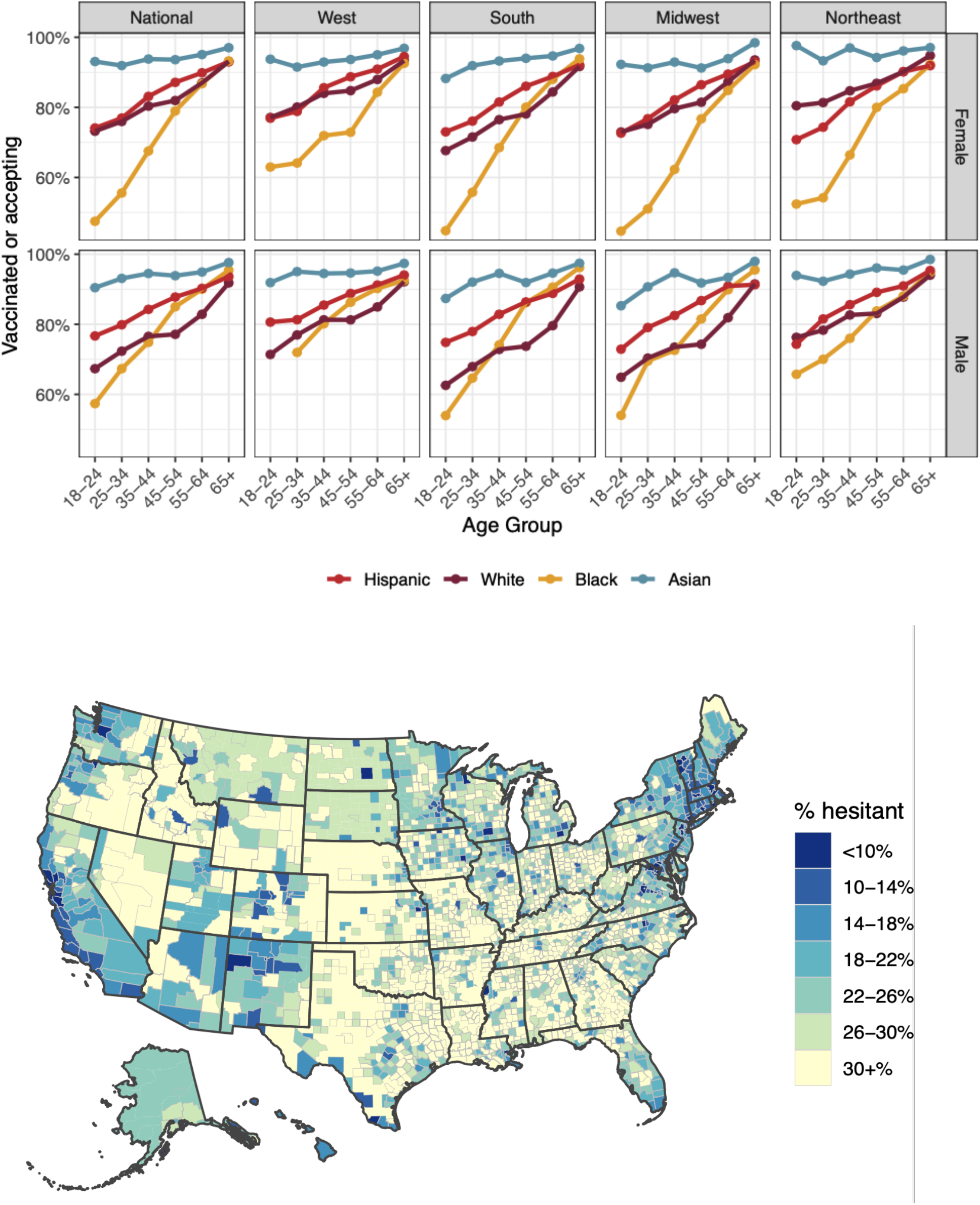
Reported vaccine acceptance and hesitancy by age group, race/ethnicity, gender, and Census region (upper panel) and by county (lower panel) during March 2021.

## Discussion

As SARS-CoV-2 spread throughout the United States during 2020 and into 2021, and policy makers faced decisions that would profoundly impact all sectors of society, the breadth and depth of information needed to support these decisions vastly exceeded the availability of data collected through existing surveillance systems designed to capture reported COVID-19 cases, hospitalizations and deaths. A number of efforts to fill the urgent need for additional information relied on novel data collection and dissemination platforms that leveraged mobile phone technology and new media. In this study, we describe one of these efforts, the COVID Trends and Impact Survey, which is the largest continuous health survey ever conducted in the United States, in operation since April 6, 2020, with more than 20 million responses collected over the first year of operation.

Comparisons to routine sources of surveillance information point to both the face validity and incremental value of the COVID Trends and Impact Survey. Time trends and geographic patterns in COVID-19 outcomes measured in CTIS – including specific symptoms strongly associated with SARS-CoV-2 infection such as anosmia, syndromic patterns such as COVID-like-illness, and the novel CTIS measure of CLI-in-community – mirror broad temporal and spatial features in standard surveillance measures on confirmed COVID-19 cases, hospitalizations and deaths, while in many cases avoiding data artifacts and reporting anomalies that affect the official measures.

In this study we have highlighted several examples of how attributes of CTIS give it particular value and salience as an information platform for public health policy. The scope, scale and recruitment strategy used in the survey support analysis at high geographic and temporal resolution, allowing detection of local trends on short timescales not available through other surveys, and accommodate a high level of stratification on relevant individual characteristics. Several examples illustrate the benefits of this granularity, including the ability to compare risks and preventive behaviors by occupational category, with further stratification possible by demography and geographic location; ability to describe variation in intentions and use of key mitigating measures including physical distancing, masking and vaccination. Regular updating of the survey has enabled the survey content to adapt alongside the evolving policy response, for example through addition of survey items on mask use in September 2020, school mitigation strategies in November 2020, and vaccination in December 2020.

Other studies have used data from the COVID Trends and Impact Survey to answer specific questions about key COVID-19 impacts and policies. A number of studies have analyzed relationships between reported risk-mitigating behaviors in the CTIS and other outcomes. For example, Reberio and colleagues (11) examined reported mask-wearing behavior as an outcome in relation to statewide mask-wearing requirements. Rader and colleagues (12) examined the relationship between mask-wearing and physical distancing as measured in CTIS and measures of community transmission. Bilinski et al. (13) assessed trends across states in a number of indicators on risk perception and preventive behaviors in relation to COVID-19 case rates. Other studies have used CTIS measures to explore correlates of variation in risk. For example, Flaxman et al. (14) computed relative infection rates for healthcare workers vs. other respondents using information from the survey on occupation, testing and test results. Lessler and colleagues (15) have assessed reported risks of COVID-19-related outcomes, including COVID-like illness, anosmia or a positive COVID-19 test, in relation to whether a household includes a child who attends in-person schooling, and reported school-based mitigation measures.

Symptom measures from the survey have also been used to aid in forecasting of COVID cases and deaths. Through the COVID-19 Forecast Hub, the CDC collects standardized forecasts from dozens of teams. Rodríguez et al. (16) incorporated symptom surveillance data from CTIS into a deep learning framework for real-time forecasting. In a companion paper in this theme issue, we demonstrate that symptom surveillance data and other auxiliary data streams (such as medical insurance claims) can improve forecasting and hotspot prediction accuracy over short (1-3 week) time intervals.

Several limitations are important to note. First, because the survey uses Facebook active users as its sampling frame and because participation in the survey is strictly voluntary, respondents may not be fully representative of the U.S. population despite incorporation of survey weights, which adjust for non-response and coverage biases based on a limited number of covariates. Comparison to the American Community Survey indicates that our sample over-represents respondents who are college-educated. Research users of the survey microdata can use additional demographic or other survey variables to construct improved post-stratification adjustments to correct this for their purposes. However, any non-response biases not accounted for by Facebook’s non-response weights would be much more difficult to correct.

Additionally, many of the outcome measures related to COVID-19 are based on self-reports, which may diverge from more objective measures due to recall bias, social desirability bias, and other sources of survey bias and measurement error. On the other hand, broad comparisons of indicators such as cumulative COVID-19 diagnoses suggest that measurement of key COVID-19 outcomes are relatively robust to response biases that may be present in the sample. Ultimately, the value of such a large-scale survey is not in accuracy afforded by its sample size, since survey biases persist no matter the size of the survey; smaller surveys more carefully constructed to reduce sampling biases would likely yield more accurate estimates (17). Instead, since these survey biases are unlikely to change rapidly in time or in space, CTIS can accurately track *trends* in key signals, even if the daily point estimates are systematically biased. This is demonstrated by the strong correlations between survey estimates of CLI-in-community and reported COVID case rates, for example; while CLI-in-community is not an unbiased population estimate of COVID case rates, it nonetheless provides useful information about trends in cases. The principal value of CTIS is hence in the detailed spatial and demographic comparisons it makes possible, and in its ability to track changes continuously over time and correlate them with key outcome measures.

Although CTIS was initially designed with a relatively limited scope, including a particular focus on syndromic surveillance, its value has ultimately derived in large part from its flexibility as a surveillance platform that can be rapidly adapted to changing information needs. Running a survey of this size has involved many challenges, particularly as it expanded to include key measures of public knowledge, attitudes, and behaviors, and as public health needs evolved continuously during the pandemic. Despite these challenges, however, CTIS has provided both a valuable public information resource during a global health emergency, as well as a potential model for ongoing health surveillance needs. Similar online surveys are likely to play important roles in future epidemics and pandemics by supplementing public reporting systems with information that is difficult to gather any other way.

## Data Availability

Aggregate data from the U.S. CTIS survey are available via the COVIDcast API, and individual data are available to researchers, subject to privacy restrictions, through data use agreement.

https://cmu-delphi.github.io/delphi-epidata/symptom-survey/

## Acknowledgements

We thank Taylor Arnold, Logan Brooks, Nat DeFries, and Kathryn Mazaitis from Delphi for contributions to survey data processing systems; Adrianne Bradford, Don Burke, Samantha Chiu, Curtiss Cobb, Xiaoyi Deng, Evan MacKay, Jonathan McKay, Stephanie Perniciaro, Stanley Presser, Liz Stuart, and Anna York for critical and insightful discussions on the initial survey design; and our many data users and collaborators who suggested survey revisions and topics.

